# Using Artificial Intelligence to optimize agreement between interstitial sensors and capillary puncture in glycemic assessment and classification

**DOI:** 10.64898/2026.02.04.26345595

**Authors:** Luana Rodrigues Ecker, Neylane Araújo Cordeiro de Santana, Camila Fernandes Caldato, Cláudio Eduardo Corrêa Teixeira

## Abstract

**Introduction:** Blood glucose monitoring is essential for the management of diabetes mellitus. Continuous interstitial glucose (IG) monitoring systems are less invasive than capillary blood glucose (BG) measurements, but their agreement decreases at higher glucose levels. Artificial intelligence (AI) approaches, particularly recurrent neural networks such as long short-term memory (LSTM), have shown potential to model temporal glucose dynamics and correct inter-method discrepancies. Objective: To develop and validate an AI-based model capable of predicting capillary BG values from IG data, improving agreement between methods and enhancing glycemic status classification. Methods: This retrospective observational study analyzed 708 paired BG-IG measurements obtained from published anonymized datasets. Data preprocessing included Kalman filtering, robust normalization, temporal windowing, and class balancing via oversampling. An LSTM model with dual output was trained to perform both capillary glucose regression and glycemic status classification. Model performance was assessed using regression metrics (MAE, RMSE, R^2^), classification metrics (accuracy, F1-score), and agreement analysis (Bland-Altman). Results: The AI model substantially reduced the mean bias from +16.27 mg/dL to -2.08 mg/dL and achieved markedly narrower limits of agreement compared with raw BG-IG differences (-129.5 to +162.0 mg/dL vs. -47.3 to +43.2 mg/dL). Glycemic classification accuracy was high for hyperglycemia (94.6%), prediabetes (93.7%) and normoglycemia (100%), with lower performance observed for hypoglycemia (66.7%). Conclusion: LSTM-based AI modeling demonstrated strong capability to predict capillary BG from IG measurements and to correct inter-method discordance. These findings support the potential integration of AI-enhanced glucose estimation into clinical monitoring systems to improve therapeutic decision-making.

## Introduction

Glycemic monitoring is one of the pillars of diabetes mellitus management and is essential for preventing acute and chronic complications associated with metabolic dysregulation (American Diabetes Association, 2023). Traditionally, capillary blood glucose (BG) obtained by fingerstick testing has been the standard method for home monitoring due to its accessibility and direct accuracy. However, this approach is considered invasive and is associated with low long-term patient adherence (Charleer et al., 2018).

With advances in continuous glucose monitoring technologies, interstitial glucose (IG) sensors have become widely used because they allow real-time monitoring with less physical discomfort (Battelino et al., 2019). Nevertheless, these sensors exhibit a relatively low level of agreement with the standard method, particularly at glycemic extremes such as hyperglycemic episodes, despite showing good linear correlation with capillary blood glucose (Klonoff et al., 2020). Bland–Altman analysis is widely used to assess agreement between methods and has revealed limits of variation that compromise the clinical reliability of sensors as substitutes for fingerstick testing (Giavarina, 2015; Caldato et al., 2026).

In this context, the application of artificial intelligence (AI) emerges as a promising alternative to correct systematic biases between monitoring methods and to improve the prediction of true capillary glucose values. The use of Long Short-Term Memory (LSTM) models has proven effective in the analysis of biomedical time series, including the prediction of glycemic patterns (Contreras & Vehi, 2018; Oviedo et al., 2017).

The objective of the present study was to develop and validate an AI model capable of predicting capillary BG values from IG sensor measurements in patients with diabetes, with a focus on optimizing agreement between methods and promoting a more reliable classification of glycemic status. This approach aims to contribute to a less invasive, more accurate, and higher-adherence monitoring strategy, with potential clinical application in personalized diabetes management.

## Methods

### Study design, data sources and ethical considerations

This quantitative, observational, and retrospective investigation was conducted using secondary glycemic data obtained from previously published and fully anonymized studies (Bailey et al., 2015; Fellinger et al., 2024). One of the source datasets is publicly available under a Creative Commons Attribution–NonCommercial license, which allows noncommercial use provided appropriate credit is given (Bailey et al., 2015), while the second dataset can be accessed upon reasonable request (Fellinger et al., 2024). Detailed baseline characteristics of the study participants are reported in the original publications.

Given that all data were anonymized and derived from prior publications, the research protocol was reviewed by the institutional Ethics Committee and classified as exempt from formal ethical approval and informed consent requirements. This determination complies with Brazilian regulations established by the National Health Council (CNS) and the National Research Ethics Commission (CONEP), including CNS Resolutions No. 510/2016 and No. 674/2022, as well as CONEP Circular Letter No. 12/2023.

The analyzed datasets consisted of paired BG-IG measurements obtained using capillary fingerstick BG testing and two generations of IG monitoring systems that are widely used: FreeStyle Libre (n = 581) and FreeStyle Libre 2 (n = 127). All analysis was performed using the pooled dataset. The total of 708 paired BG-IG measurements were randomly extracted from graphical data presented in the original sources using WebPlotDigitizer software (version 4.6; Automeris, USA), a validated and widely adopted tool for numerical data extraction from scientific figures. Linear association between BG and IG measurement methods was assessed using Pearson’s correlation coefficient (r), while the coefficient of determination (R^2^) was used to estimate the proportion of BG variability explained by IG values. Agreement between methods was evaluated through Bland–Altman analysis, in which the difference between paired measurements (BG *minus* IG) was examined as a function of their mean (Giavarina, 2015).

### Data Preprocessing and Structuring

For the purposes of model development and validation, the complete dataset was partitioned into two subsets, with 90% of the data allocated to training and cross-validation and the remaining 10% reserved for independent final validation, following established best practices in deep learning research (Goodfellow et al., 2016).

To minimize noise and abrupt variations in the time series, a Kalman filter was applied, a statistical technique effective for signal smoothing and widely validated in biomedical applications (Knobbe et al., 2005). Subsequently, sliding temporal windows (lags) with shifts of 1, 5, 10, and 15 samples were generated to capture relevant temporal patterns in IG.

The data were normalized using a RobustScaler in order to mitigate the effects of outliers, which are particularly frequent in extreme glycemic measurements. The regression target variable was BG, while categorical glycemic state classification followed clinical guidelines (Rodacki et al., 2024): hypoglycemia < 70 mg/dL; normoglycemia 70–100 mg/dL; prediabetes 101–126 mg/dL; and hyperglycemia > 126 mg/dL. Given the natural class imbalance, manual oversampling of minority classes was applied, an effective practice to prevent bias in multiclass classifiers (Chawla et al., 2002).

### Neural Network Architecture and Training Strategy

A model based on Recurrent Neural Networks (RNN) with a Long Short-Term Memory (LSTM) architecture was implemented, given its recognized ability to capture long-term dependencies in physiological time-series data (Sun et al., 2018). The model was structured with two sequential LSTM layers (64 and 32 units), interleaved with Dropout layers (30%) to prevent overfitting (Srivastava et al., 2014).

The model included two parallel outputs: a continuous regression output with linear activation to predict numerical BG values, and a multiclass classification output with softmax activation to classify glycemic state. The composite loss function included mean absolute error (MAE) for regression and weighted cross-entropy for classification, with balanced weights of 0.5 for each component. Training was performed using the Adam optimizer, with early stopping based on validation loss, as recommended for robust deep network training (Kingma & Ba, 2014).

### Performance Evaluation and Visualization

For regression output evaluation, MAE (Mean Absolute Error), RMSE (Root Mean Square Error), and R^2^ (Coefficient of Determination) were calculated. Classification performance was assessed using overall accuracy, confusion matrices, and classification reports (precision, recall, and F1-score). In addition, agreement between true BG and predicted BG values was evaluated using Bland–Altman analysis. Scatter plots, training loss curves, and confusion matrices were also generated to visualize predictive performance.

## Results

The initial analysis of the linear correlation between BG and IG values revealed a strong positive association (r = 0.88), as illustrated in Figure 1 (left). However, a substantial dispersion of data points around the line of perfect correlation was observed, indicating that, despite being high, correlation alone does not ensure agreement between the two glucose measurement methods.

**Figure 1.**
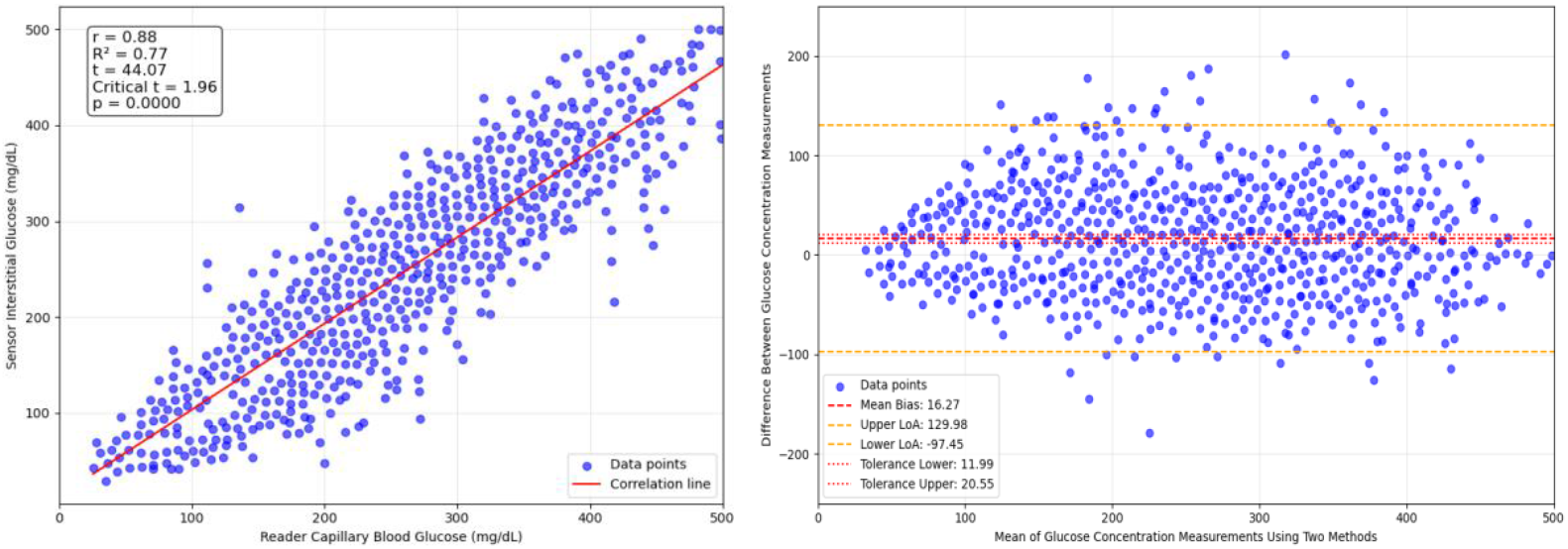
Left: Linear correlation between capillary BG and IG values. A strong correlation is observed (r = 0.88), but with high dispersion of data points around the regression line, indicating low individual precision. Right: Bland–Altman plot comparing BG and IG. The positive mean bias demonstrates systematic underestimation of IG relative to BG, with wide limits of agreement.

In Figure 1 (right), agreement analysis between BG and IG using the Bland–Altman plot showed a mean bias of +16.27 mg/dL, indicating that IG sensors tend to underestimate true BG values. In addition, the limits of agreement ranged from –129.5 to +162.0 mg/dL, demonstrating a wide margin of error that compromises the clinical accuracy of the sensor as a substitute for fingerstick testing.

In contrast, capillary BG prediction using AI demonstrated substantially superior results. Figure 2 (left) shows the scatter plot between actual BG values and BG values predicted from IG values by the AI model, with data points distributed close to the line of perfect prediction. It is evident that the correlation between these variables (r = 0.97) is better than that observed between BG and IG values in Figure 1 (left).

**Figure 2.**
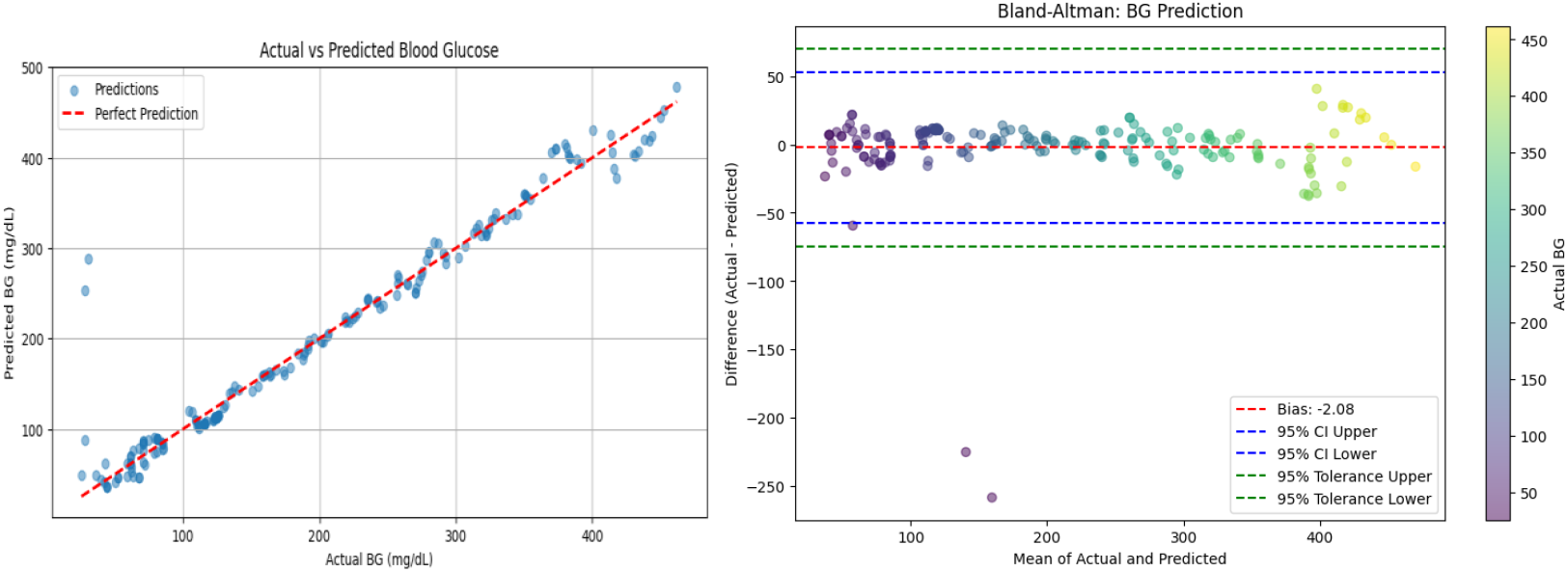
Left: Linear correlation between actual capillary BG values and AI-predicted BG. A very strong correlation is observed (r = 0.97), with low dispersion of data points around the regression line, indicating high individual precision. Right: Bland-Altman plot comparing BG and AI-predicted BG. The reduced mean bias and narrower limits of agreement indicate improved concordance between glucose values obtained by these methods.

However, Figure 2 (right) shows what is more relevant than higher correlation when comparing BG and IG measurements: the Bland–Altman plot comparing actual BG and predicted BG values, showing a lower mean bias of -2.08 mg/dL, with limits of agreement also significantly narrower (-47.3 to +43.2 mg/dL), highlighting a substantial improvement in prediction reliability compared with the original IG measurement method.

During neural network training, a progressive reduction in both glycemic prediction errors (BG Loss) and glycemic state classification errors (Class Loss) was observed, as shown in Figure 3, with MAE = 12.29, RMSE = 28.06, accuracy = 0.91, and F1-scores of 0.80 (hypoglycemia), 0.82 (normoglycemia), 0.88 (prediabetes), and 0.96 (hyperglycemia). Both training and validation sets exhibited decreasing loss curves, suggesting model convergence and absence of overfitting.

**Figure 3.**
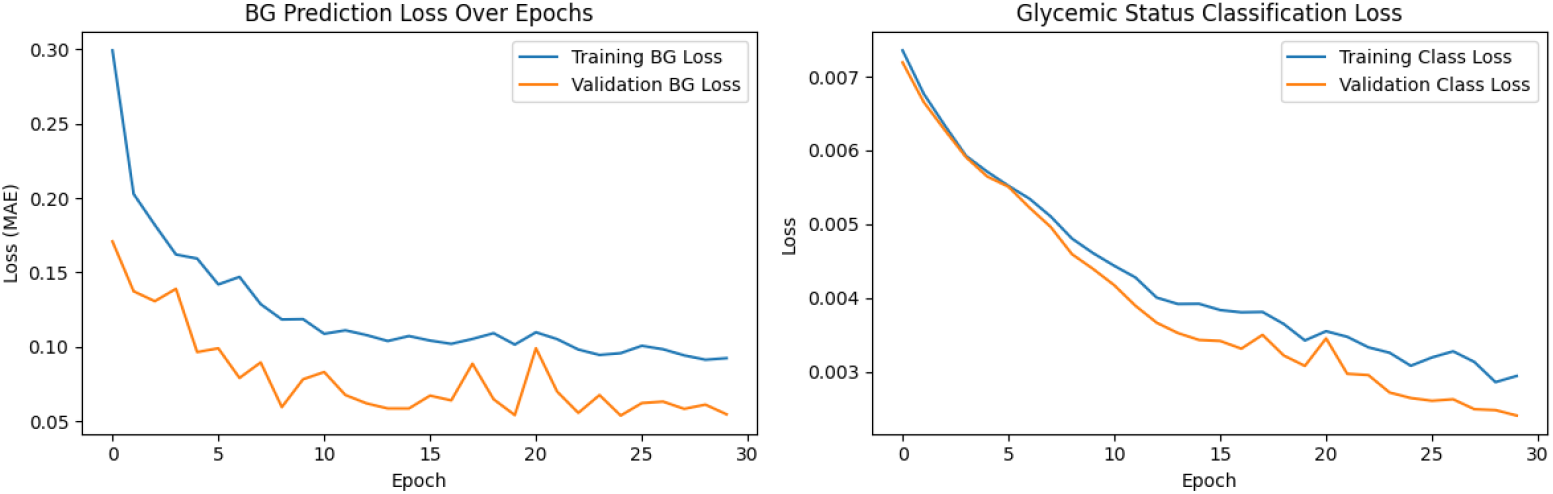
Loss curves during neural network training. Left: reduction in glucose prediction loss (MAE). Right: reduction in glycemic state classification loss, for both training and validation data.

Finally, Figure 4 presents the confusion matrix of the AI model for glycemic state classification. High accuracy is observed in absolute numbers for hyperglycemia (94.6%), prediabetes (93.7%) and normoglycemia (100%), along with satisfactory performance for hypoglycemia (66.7%). Errors occurred mainly in the classification of hypoglycemic cases as normoglycemic, suggesting the need for further refinement of the model in this glycemic range.

**Figure 4.**
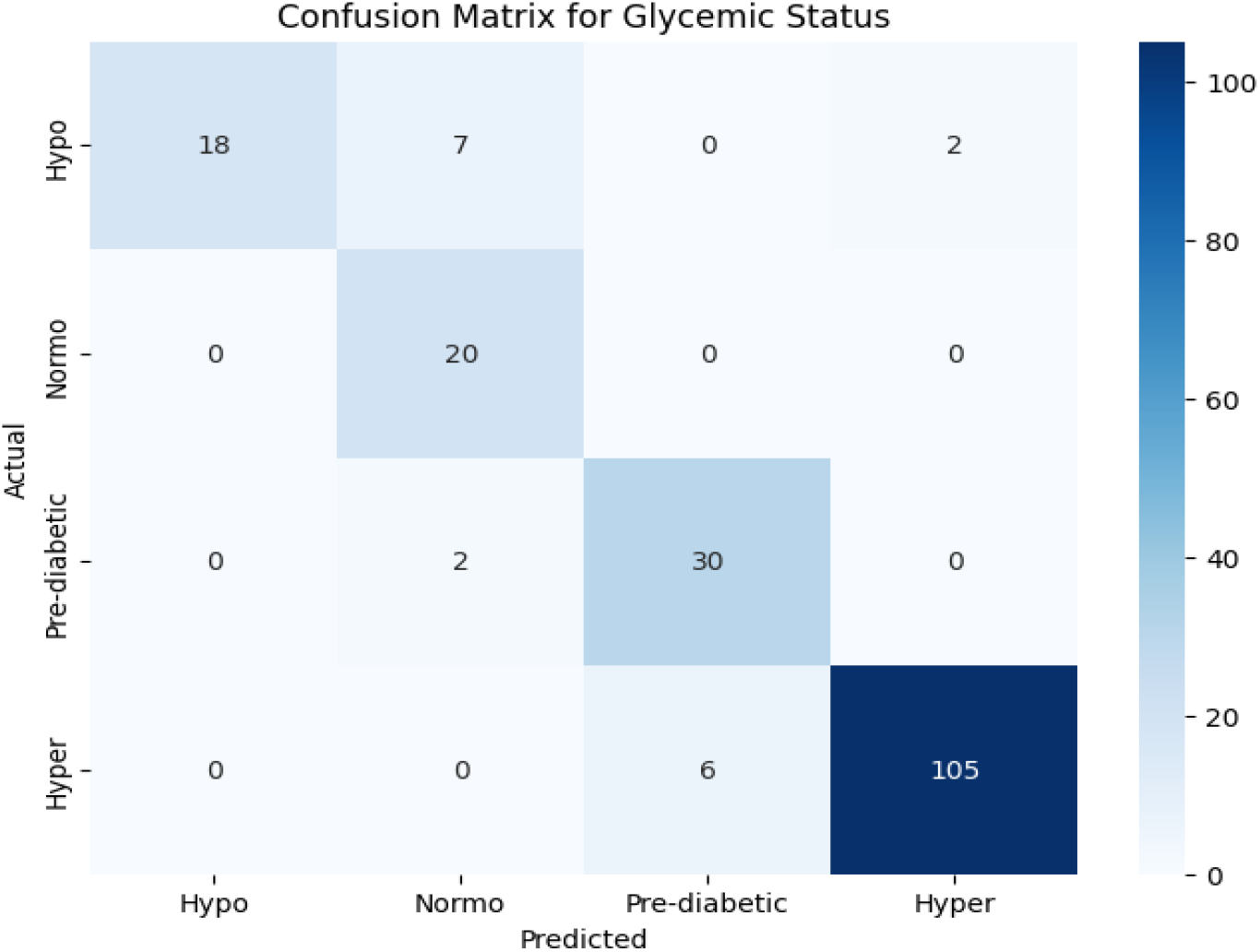
Confusion matrix for AI-based glycemic state classification. The AI achieved superior performance in the hyperglycemia, prediabetes and normoglycemia categories.

## Discussion

The use of AI for predicting glycemic parameters has gained increasing attention as a promising strategy to optimize the management of diabetes mellitus, particularly in patients who rely on IG sensors for daily disease monitoring. In the present study, we demonstrated that an LSTM-based neural network model was able to significantly improve agreement between values estimated by interstitial sensors and true values obtained by capillary fingerstick testing, overcoming limitations previously described in the literature.

### Discrepancy between sensors and capillary fingerstick testing: a clinically relevant problem

Although interstitial sensors are recommended by international societies for continuous glucose monitoring due to their convenience and lower invasiveness, substantial evidence indicates that these devices do not ensure satisfactory clinical agreement with capillary BG, especially at glycemic extremes (hypo- and hyperglycemia) (Klonoff et al., 2020; Heinemann, 2018). The Bland–Altman analysis performed in this study confirms this finding, revealing a significant positive bias and wide limits of agreement, thereby reinforcing the need for adjustments to achieve full clinical reliability.

### AI performance in correcting discrepancies and predicting glycemia

The application of AI, particularly LSTM-based models, proved effective in markedly reducing the mean bias and dispersion observed in Bland-Altman analyses between actual and predicted values. This performance is consistent with previous studies that employed deep neural networks for glycemic prediction, such as those by Oviedo et al. (2017) and Li et al. (2019), which demonstrated that LSTM is among the most effective architectures for capturing temporal patterns in glucose time series.

In addition, glycemic state classification accuracy was high, especially for normoglycemic, prediabetes and hyperglycemic states. The lower accuracy observed for hypoglycemic episodes is consistent with the literature, which suggests that such events are more difficult to predict due to their rapid and unstable nature (Zecchin et al., 2016). Nevertheless, performance levels remained superior to those reported for traditional methods or standalone commercial sensors.

### Clinical implications and future perspectives

The potential use of AI as a support tool to correct inaccurate sensors readings has direct implications for clinical practice. Models such as the one proposed in this study could be embedded in wearable devices or mobile applications, functioning as real-time verification or correction systems, thereby enhancing therapeutic safety and enabling more precise insulin therapy adjustments (Contreras & Vehi, 2018).

Another potential benefit is the reduction in the need for frequent capillary fingerstick testing. Studies indicate that pain and discomfort associated with fingerstick procedures are major contributors to poor adherence to traditional glycemic monitoring, particularly among younger populations or individuals with needle phobia (Russell-Jones et al., 2018). Thus, a validated AI model may promote greater treatment adherence, reduce adverse events, and improve long-term glycemic control.

### Limitations and methodological considerations

Despite the promising results, this study has some limitations. The dataset was extracted from published graphs, which may limit the generalizability of the findings to more heterogeneous populations. In addition, model performance may vary depending on sensor quality, measurement frequency, and patient clinical profile.

For future studies, we suggest the use of prospective clinical cohorts, comparison with other AI architectures (such as transformers or hybrid models), and external validation of the model in diverse populations, including individuals with type 1 diabetes, type 2 diabetes, and gestational diabetes.

## Conclusion

This study demonstrated that the application of AI-based models, specifically recurrent neural networks of the LSTM type, is an effective strategy to enhance agreement between IG sensors and BG values obtained through capillary fingerstick testing. The AI model was able to significantly reduce bias and narrow the limits of agreement compared with commercial sensors, while also achieving high accuracy in glycemic state classification.

These findings reinforce the clinical potential of AI as a supportive tool for glycemic monitoring, enabling a less invasive, more accurate, and more adaptable approach tailored to the individual needs of patients with diabetes. The integration of such predictive systems into continuous-use devices may improve treatment adherence, enhance the safety of therapeutic decisions, and improve patients’ quality of life.

Nevertheless, future studies are recommended to include larger and more diverse clinical samples, multicenter validation, and comparisons with different machine learning architectures. Such advances will help establish AI as a fundamental component of personalized diabetes mellitus management.

## Data Availability

All data produced in the present work are contained in the manuscript.

